# Healthcare use in individuals with rheumatoid arthritis during the COVID-19 pandemic and beyond: A cohort study in three nations of the UK

**DOI:** 10.1101/2025.04.25.25326408

**Authors:** Ruth E Costello, Michael Parker, Jonathan Kennedy, Sinead Brophy, Amir Mehrkar, Sebastian Bacon, Ben Goldacre, Brian MacKenna, Dave Evans, Laurie Tomlinson, Rosemary Hollick, Jenny H Humphreys, the OpenSAFELY collective, the RHEUMAP’s study investigators and the LH&W NCS (or CONVALESCENCE) Collaborative

**Affiliations:** London School of Hygiene and Tropical Medicine, Keppel Street, London WC1E 7HT, UK; National Centre for Population Health and Wellbeing Research, Swansea University Medical School, Wales, SA2 8PP; Bennett Institute for Applied Data Science, Nuffield Department of Primary Care Health Sciences, University of Oxford, Oxford, UK; Aberdeen Centre for Arthritis and Musculoskeletal Health (Epidemiology Group), University of Aberdeen, Health Sciences Building, Foresterhill Campus, Aberdeen AB25 2ZD; Centre for Epidemiology Versus Arthritis, University of Manchester, Manchester Academic Health Centre, Manchester, UK

**Keywords:** Rheumatoid Arthritis, delivery of health care, inequalities, observational studies, organisation of health care

## Abstract

**Objectives:** We aimed to estimate how rheumatology healthcare use has changed since the COVID-19 pandemic and determine demographic characteristics associated with observed changes in healthcare use.

**Methods:** Using three primary and secondary care electronic health record datasets in England (with the approval of NHS England), Scotland, and Wales, we identified individuals with a diagnosis of rheumatoid arthritis (RA) before 01/04/2019. We determined the proportion of people with rheumatology hospital outpatient appointments each month (April 2019-December 2022 (Wales and Scotland), April 2019-November 2023 (England)) and quantified changes using interrupted time-series analysis. We used logistic regression to determine characteristics associated with having fewer appointments compared to 2019.

**Results:** We identified 145,065, 3,813 and 13,637 individuals coded with RA in England, Scotland, and Wales, respectively. At the start of the COVID-19 pandemic the number of rheumatology outpatient appointments dropped sharply across all nations. In England and Scotland, the percentage of monthly appointments has continued to decline. In Wales, while there was a gradual recovery, rheumatology services have not returned to pre-pandemic levels. In contrast, the number of appointments for all other specialist outpatient appointments have recovered in all nations. Ethnic minorities, those living in more deprived areas and urban areas had fewer appointments after the start of the pandemic compared to 2019.

**Conclusion:** For the first time, we compared healthcare use across three UK nations and found that rheumatology outpatient appointments had not recovered to pre-COVID-19 pandemic levels, particularly in Scotland and England. Certain patient groups had fewer appointments during the study period.

**Key messages:**

- Rheumatology outpatient appointments remain below pre-pandemic levels, particularly in England and Scotland, unlike other specialties.
- Ethnic minorities, deprived communities, and urban residents had fewer rheumatology appointments post-pandemic than in 2019. Rheumatology services need data-driven strategies to provide better support, tailored to local community needs.

## Introduction

Rheumatoid arthritis (RA) is a chronic inflammatory condition that requires ongoing specialist care. Rheumatologists employ a treat-to-target strategy, with the goal of keeping disease activity low, with frequent monitoring and adaptation of treatments to reach this goal (1). To achieve this in early inflammatory arthritis, monitoring every one to three months is recommended (2). In the UK this can move to annual review once the disease is stable. This enables monitoring of disease activity and the development of comorbidities. Patients should have access to specialist care in the case of disease flare-ups (3).

Rheumatology services were significantly affected by the COVID-19 pandemic, with rheumatologists frequently seconded to help with treating COVID-19 patients. Rheumatology and primary care appointments were moved to telephone or video consultations and drug monitoring was reduced (4,5). It is not clear how this impacted people with RA. There have been reports of medication interruptions during the pandemic, resulting in disease flares.

However, many of these studies are based on self-reported data (6–8). Beyond the impact of the pandemic, health inequities exist in RA, with lower deprivation being associated with worse outcomes (9,10). There is evidence that individuals of lower socioeconomic status (SES), older age and those living in a rural location can struggle to access rheumatology services (11).

Prior to the COVID-19 pandemic, several countries have reported that there is a shortage of rheumatologists, which is predicted to worsen (4,12,13). The UK rheumatology workforce is understaffed. It is recommended that there is one consultant per 60,000 population, however, there is significant variation across the devolved nations. For example, there is one consultant per 80,617 in England and one per 111,637 in Scotland, resulting in potential gaps in care (4).

Furthermore, in the UK, healthcare is delivered under the umbrella of the National Health Service (NHS), however health is devolved across the four nations of the UK (England, Wales, Scotland, and Northern Ireland). Each nation has variation within healthcare systems, with diverse policies and priorities.

We had access to linked primary and secondary electronic health record (EHR) data from three of the four nations (England, Wales, and Scotland) providing a unique opportunity to explore healthcare use across devolved nations of the UK and examine the differential impacts since the COVID-19 pandemic. In patients with RA we aimed to 1) estimate how hospital outpatient appointments have changed over the course of the pandemic, compared to 2019, and 2) to determine socio-demographic characteristics associated with any observed changes in healthcare use.

## Methods

### Study Design and Data Sources

We conducted a population-based observational cohort study of people with RA using electronic health records (EHR) in England, Wales, and Scotland.

In England, we used primary care records managed by the GP software provider TPP linked to outpatient appointment data from the Hospital Episode Statistics for England and mortality data from the Office for National Statistics (ONS) through OpenSAFELY. TPP patients represent 42.5% of the UK population and are broadly representative (14). All data were linked, stored, and analysed securely within the OpenSAFELY platform: https://opensafely.org/, as part of the NHS England OpenSAFELY COVID-19 service. Data include pseudonymised data such as coded diagnoses, medications, and physiological parameters. No free text data are included. All code is shared openly for review and re-use under MIT open licence [https://github.com/opensafely/RA_outcomes]. Detailed pseudonymised patient data is potentially re-identifiable and therefore not shared.

Primary care data for 85% of the Welsh population is available within the SAIL Databank. All people alive in Wales registered with a general practice who contribute data to SAIL were identified as of 23^rd^ March 2020. Individuals with diagnostic codes for RA from 1^st^ January 2005 to 22^nd^ March 2020 were identified from this general population group using primary care records in the Welsh Longitudinal General Practice (WLGP) database. This information was linked to other national databases in SAIL including: outpatient appointments, emergency care and hospital admissions (15).

In contrast, there is no national, anonymised primary care dataset in Scotland and primary care data can only be accessed through a trusted third-party provider. This process currently requires written permission from individual GP practices and reimbursement for time to complete the agreements. Due to time and financial constraints, this practically limited the scope of data collection in Scotland. We report on primary care data from two health boards in Scotland: Grampian and Highland. These were selected to provide a mix of urban, accessible, and remote rural mainland communities and island communities and different healthcare settings. The data presented here are limited to practices that granted permission, representing 50% of practices approached. Primary care data was linked to other national databases in Public Health Scotland, including outpatient hospital appointments, hospital admission records, registries for death and cancer, and all medications dispensed in community care. Data linkage was conducted by the NHS Scotland electronic Data Research and Innovation Service via deterministic linkage methods using unique personal identification numbers in a process shown to produce highly accurate and complete data. Data was accessed with the National Data Safehaven.

### Study Population

We identified people with coded RA on or prior to 1st April 2019 (index date) using a validated algorithm (16). The algorithm uses diagnosis (Read or SNOMED CT codes) and prescription codes from the primary care EHRs. In England and Wales people were considered to have a diagnosis of RA if they had either 1) ≥2 RA diagnosis codes on different dates, or 2) a single RA diagnosis code and disease-modifying anti-rheumatic drug (DMARD) prescribing after the first RA code. In Scotland, due to lack of sufficient medication data, diagnosis was based on >=2 RA codes. We required people to have at least 3 months registration with their GP prior to 1st April 2019 and be aged 18-115 years. We excluded people with missing age, sex, region, or deprivation as this could indicate poor data quality. We followed people until the earliest of death, deregistration from their practice or the end of the study period (Wales and Scotland: 31st December 2022, England: 30^th^ November 2023).

### Study Measures

Using secondary care data, we identified all hospital outpatient appointments within the study period and then classified appointments by whether they were with the rheumatology specialty or other medical and surgical specialties. In England these were appointments attended, in Scotland and Wales, these were appointments scheduled.

We determined socio-demographic characteristics at index date including age, sex, ethnicity, urban-rural location of residence, and deprivation (Table 1) (17–19). We identified smoking status at index date by identifying the latest smoking code and any record of prior smoking, and classified people as either non-smokers, ex-smokers or current smokers. BMI at baseline was defined using the most recent measurement within the 5 years prior to baseline.

**Table 1.** Covariate definitions for each nation.

|  | England | Wales | Scotland |
| --- | --- | --- | --- |
| Region | 9 regions of England based on NHS administrative geographical areas | 22 Local Authorities | 2 Health boards (Highlands and Grampian) |
| Rural-urban classification | Based on Rural Urban Classification for England and Wales (20) and categorised into urban and rural. | Based on Rural Urban Classification for England and Wales (20) and categorised into urban and rural. | Based on Rural Urban Classification for Scotland (21) and categorised into urban and rural. |
| Deprivation | Quintiles of Index of Multiple Deprivation (IMD) based on a person's postcode (17). | Quintiles based on the Welsh Index of Multiple Deprivation (WIMD) based on 1990 LSOAs (18). | Quintiles based on the Scottish Index of Deprivation (SIMD) based on 6976 data zones (19). |
| Ethnicity | Self-reported ethnicity identified using SNOMED CT codes in primary care record, supplemented with ethnicity recorded in the secondary care record. | Not available | Not available |

### Statistical Analysis

We describe the baseline socio-demographic characteristics and healthcare use of each nation. For outpatient appointments with rheumatology and all other hospital-based specialties, we determined the proportion of people who attended/scheduled appointments each month. We quantify changes using time-series analysis, where monthly proportions were modelled in an ordinary least-squares regression model with Newey-West heteroskedasticity-consistent standard errors and one lag to account for autocorrelation. The interruption was set at 23rd March 2020. To further understand how appointments changed over time, and because frequently people are seen on an annual basis in rheumatology, we counted the number of outpatient appointments with 1) rheumatology and 2) all other specialties each year: April - March. People were classified as having either zero, 1-2 or ≥3 rheumatology outpatient appointments in a given year. People were included in the year count if they were followed for the whole year. We compared the COVID-19 pandemic year counts (years starting April 2020 onwards) to the year prior to the pandemic (year starting April 2019) and determined if people had either no appointments in either year, fewer appointments, or the same number or more appointments since the start of the COVID-19 pandemic. In people who had appointments in both years, we describe the characteristics of these groups and used logistic regression to measure whether age, sex, ethnicity, rural-urban classification, area-based measure of deprivation and region were associated with having fewer appointments, as this could represent groups not receiving equitable care. We modelled the characteristics individually in univariate models, and in a multivariate model and we present the odds ratio (OR) and 95% confidence interval (CI) for having fewer appointments compared to 2019.

### Software and Reproducibility

Data management in England was performed using Python 3.9.7, with analysis carried out using Stata 17. Code for data management and analysis, as well as codelists and the protocol, are archived online [https://github.com/opensafely/RA_outcomes/tree/main].

Data management in Wales was primarily conducted on an IBM database (DB2) using SQL. Scottish data was received as comma-separated-value files which were imported into RStudio 4.4.1 for handling. In Wales and Scotland analysis performed using RStudio 4.4.1. Data visualisations combining all nation data were also created in RStudio using ggplot2 and other open-source libraries.

## Results

### Demographics

The number of people with RA on 1^st^ April 2019 in the three nation cohorts was as follows;145,065 (England), 3,813 (Scotland) and 13,637 (Wales). The English Cohort was slightly older (Age >80 years: 15.9% (England), 11.7% (Wales), and 12.67% (Scotland)), and less rural than Scotland or Wales (Rural: 24.7% (England), 32.0% (Wales), and 33.7% (Scotland)). The Scottish cohort was less deprived than England and Wales (IMD 1 (most deprived): 17.9% (England), 19.3% (Wales), 6.4% Scotland)). The English cohort had a longer disease duration at study start than Wales and Scotland (Time since first RA code: 12.2 years (England), 9.5 years (Wales), 7.6 years (Scotland)) (Table 2).

**Table 2.** Characteristics of each cohort at study start (1st April 2019)

|  |  | England* | Wales | Scotland |
| --- | --- | --- | --- | --- |
| N |  | N=145,065 | N=13,637 | N=3,813 |
| Age, n (%) | 18-40 years | 7910 (5.5) | 775 (5.68) | 279 (7.32) |
|  | 41-60 years | 39305 (27.1) | 4055 (29.74) | 1174 (30.79) |
|  | 61-80 years | 74805 (51.6) | 7258 (53.22) | 1877 (49.23) |
|  | 80+ years | 23045 (15.9) | 1549 (11.36) | 483 (12.67) |
| Sex, n (%) | Female | 102145 (70.4) | 9533 (69.91) | 2557 (67.06) |
|  | Male | 42925 (29.6) | 4104 (30.09) | 1256 (32.94) |
| Rural-Urban classification, n (%) | Rural | 35825 (24.7) | 4362 (31.99) | 2527 (66.27) |
|  | Urban | 109245 (75.3) | 9275 (68.01) | 1286 (33.73) |
| Index of Multiple Deprivation, n (%) | 1 (most deprived) | 25995 (17.9) | 2637 (19.34) | 245 (6.43) |
|  | 2 | 28310 (19.5) | 3037 (22.27) | 601 (15.76) |
|  | 3 | 32695 (22.5) | 2871 (21.05) | 1046 (27.43) |
|  | 4 | 30375 (20.9) | 2605 (19.1) | 1338 (35.09) |
|  | 5 (Least deprived) | 27695 (19.1) | 2487 (18.24) | 583 (15.29) |
| Smoking, n (%) | Never | 53555 (36.9) | 3496 (25.64) | - |
|  | Current | 20390 (14.1) | 3108 (22.79) | - |
|  | Former | 70740 (48.8) | 7033 (51.57) | - |
|  | Unknown | 380 (0.3) | - | - |
| BMI, n (%) | Underweight | 3605 (2.5) | - | - |
|  | Healthy weight | 38440 (26.5) | - | - |
|  | Overweight | 42120 (29) | - | - |
|  | Obese | 34375 (23.7) | - | - |
|  | Severe obesity | 6250 (4.3) | - | - |
|  | Unknown | 20280 (14) | - | - |
| Ethnicity, n (%) | White | 131975 (91) | 6227 (45.66) | - |
|  | Asian | 8210 (5.7) | - | - |
|  | Black | 1910 (1.3) | - | - |
|  | Mixed | 890 (0.6) | - | - |
|  | Other | 1085 (0.7) | - | - |
|  | Non-White |  | 78 (0.57) | - |
|  | Missing | 1000 (0.7) | 7332 (53.77) | - |
| Time since first RA code (years), mean (SD) |  | 12.21 (10.97) | 9.49 (5.63) | 7.63 (5.01) |

\* Counts for England rounded to the nearest 5.

### Outpatient Hospital Appointments Per Month

In England and Wales, in the year prior to the COVID-19 pandemic on average 14% of the cohorts attended a rheumatology outpatient appointment each month. In Scotland 8% of the cohort attended a rheumatology outpatient appointment each month. Across all nations there was a marked reduction in the proportion of patients having rheumatology appointments at the start of the COVID-19 pandemic (March 2020).

In Wales, the proportion of the cohort with a rheumatology outpatients appointment increased from 12% in June 2020 to 13% in August 2022. In Scotland and England during the same period, the proportion with a rheumatology appointment each month decreased over time from 5.4% to 4.2% in Scotland, and from 13% to 11% in England (Figure 1). For other speciality outpatient appointments, although there was a similar reduction in appointments at the start of the COVID-19 pandemic, appointments recovered to at least pre-pandemic levels in all nations (Figure 1).

**Figure 1.**
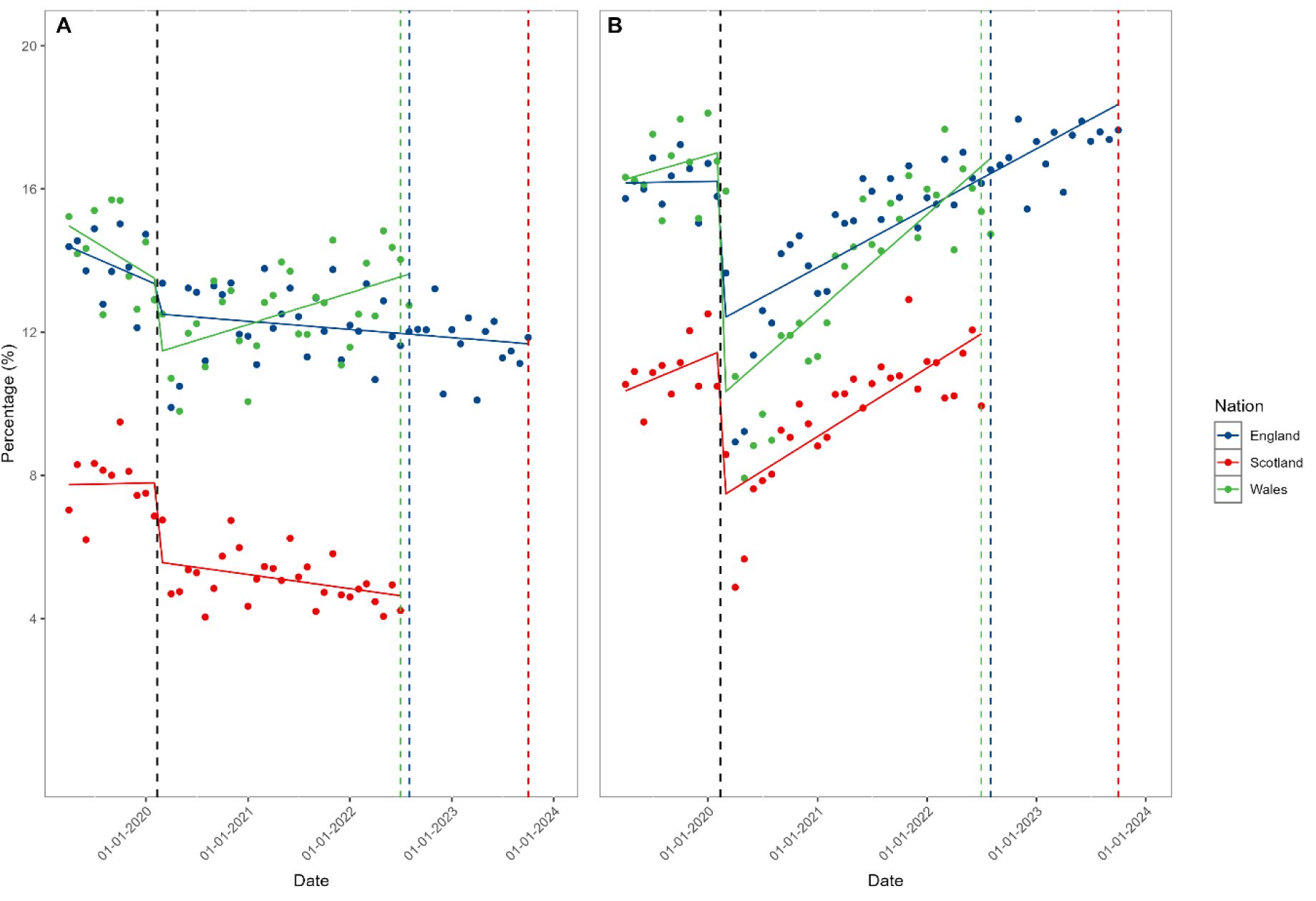
Percentage of the cohorts to attending (England)/scheduled to attend (Scotland/Wales) rheumatology (A) or all other (B) outpatient appointments each month. Black dotted line indicates the beginning of lockdown restrictions in the UK. Coloured dotted lines indicate the end of the study period in each nation.

### Yearly outpatient hospital appointments

England and Wales had similar patterns in the number of patients attending or scheduled to attend appointments per year. In the year prior to the COVID-19 pandemic (April 2019-March2020), people most frequently had 1-2 rheumatology outpatient appointments per year (England: 44%, Wales: 46%), with around one-third having no appointments (England: 30%, Wales: 27%) and a quarter having 3 or more appointments per year (England: 25%, Wales 27%). This pattern continued after the start of the COVID-19 pandemic, however there was a higher proportion of people who had no appointments (year starting April 2020: 33.6% (England), 32.5% (Wales)). When comparing 2020 to 2019, compared to Wales; a higher proportion of patients in England had no rheumatology appointments in either year (22% England vs. 18% Wales). In Wales, 38% patients had fewer appointments (compared to 35% in England).

In Scotland, in the year prior to the COVID-19 pandemic, 47% of people had no rheumatology outpatient appointments, 44% had 1-2 appointments and 9% had 3 or more appointments. In the year after the COVID-19 pandemic, the proportion of people with no appointments increased to 63%. When comparing 2020 to 2019, 41% had no appointments in either year, and 33% had fewer appointments (Table 3).

**Table 3.** Number of rheumatology outpatient appointments in comparison to 2019, across each nation and each year.

|  |  | April 2019-March 2020 |  |  | April 2020 – March 2021 |  |  | April 2021 – March 2022 |  |  | April 2022 - March 2023 |  |  |
| --- | --- | --- | --- | --- | --- | --- | --- | --- | --- | --- | --- | --- | --- |
|  |  | England | Wales | Scotland | England | Wales | Scotland | England | Wales | Scotland | England | Wales | Scotland |
| N* |  | 138,835 | 13,021 | 3614 | 132,085 | 12,351 | 3412 | 125,640 | 11,791 | 3238 | 119,295 | - | - |
| Number of rheumatology appointments, n (%) | Zero appointments | 42,180 (30.4) | 3,498 (26.86) | 1705 (47.18) | 44,430 (33.6) | 4,014 (32.5) | 2146 (62.9) | 42,450 (33.8) | 4,204 (35.65) | 2135 (65.94) | 41,245 (34.6) | - | - |
|  | 1-2 per year | 61,705 (44.4) | 5,947 (45.67) | 1567 (43.36) | 59,160 (44.8) | 5,350 (43.32) | 1039 (30.45) | 55,360 (44.1) | 4,441 (37.66) | 852 (26.31) | 53,210 (44.6) | - | - |
|  | 3 or more per year | 34,950 (25.2) | 3,576 (27.46) | 342 (9.46) | 28,495 (21.6) | 2,987 (24.18) | 227 (6.65) | 27,830 (22.2) | 3,146 (26.68) | 251 (7.75) | 24,840 (20.8) | - | - |
| Difference in number of rheumatology appointments compared to 2019, n (%) | Zero appointments both years | - | - | - | 28,905 (21.9) | 2,271 (18.4) | 1411 (41.35) | 26,490 (21.1) | 2,127 (17.22) | 1356 (41.88) | 24,245 (20.3) | - | - |
|  | Fewer appointments | - | - | - | 45,555 (34.5) | 4,666 (37.8) | 1141 (33.44) | 44,675 (35.6) | 4,461 (36.12) | 1106 (34.16) | 45,235 (37.9) | - | - |
|  | Same number or more appointments | - | - | - | 57,625 (43.6) | 5,414 (43.8) | 860 (25.21) | 54,480 (43.4) | 5,203 (42.13) | 776 (23.97) | 25,570 (41.8) | - | - |

\* Counts for England rounded to the nearest 5.

For non-rheumatology specialists’ outpatient appointments, in the year prior to the COVID-19 pandemic, a lower proportion of people had no appointments ((14.8% (England), 12.4% (Wales) and 25.8% (Scotland)) compared to rheumatology. In England and Wales, people frequently had over 3 non-rheumatology appointments per year. In Scotland people most frequently had 1-2 appointments. When compared to 2019, people frequently had fewer appointments but less frequently had no appointments in either year in all nations in contrast to rheumatology (Table S1, supplementary materials).

### Differences in healthcare use by sociodemographic characteristics

Across all nations, those with no scheduled or attended outpatient hospital appointments were older than 80 years, more frequently male and were more likely to live rurally (Table S2 & S3, supplementary materials).

For those with outpatient hospital appointments, comparing 2020 to 2019, in England, those in the least deprived quintiles (3–5) less likely to have fewer appointments compared to the most deprived quintile (IMD 5: OR 0.94, 95% CI 0.90 to 0.98, IMD 4: OR 0.94, 95% CI 0.90 to 0.98. IMD 3: OR 0.95, 95% CI 0.91 to 0.99) (Figure 2, Table S4 supplementary materials). A similar pattern was seen in Wales and Scotland, though with fewer individuals, there was greater uncertainty around the estimates. In Wales and Scotland, those living in an urban area had fewer appointments (OR (95% C)I: 1.19 (1.04, 1.35) (Wales), 1.70 (1.44, 2.00) (Scotland)), there was little difference in England (OR 95% CI: 1.02 (0.99, 1.05)). In Wales and Scotland, males were less likely to have fewer appointments (OR 95% CI: 0.83 (0.74, 0.94) (Wales), 0.81 (0.69, 0.95) (Scotland)), but this was not seen in England (OR 95% CI: 0.99 (0.96, 1.01)). In England, those with Mixed, Asian or Other ethnicity had fewer appointments (Asian: OR 1.21, 95% CI 1.17, 1.28, Mixed: OR 1.26, 95% CI 1.08, 1.47, Other: OR 1.18, 95% CI 1.03, 1.37). Similar patterns were seen when comparing 2021to 2019 (Table S5, supplementary materials).

**Figure 2:**
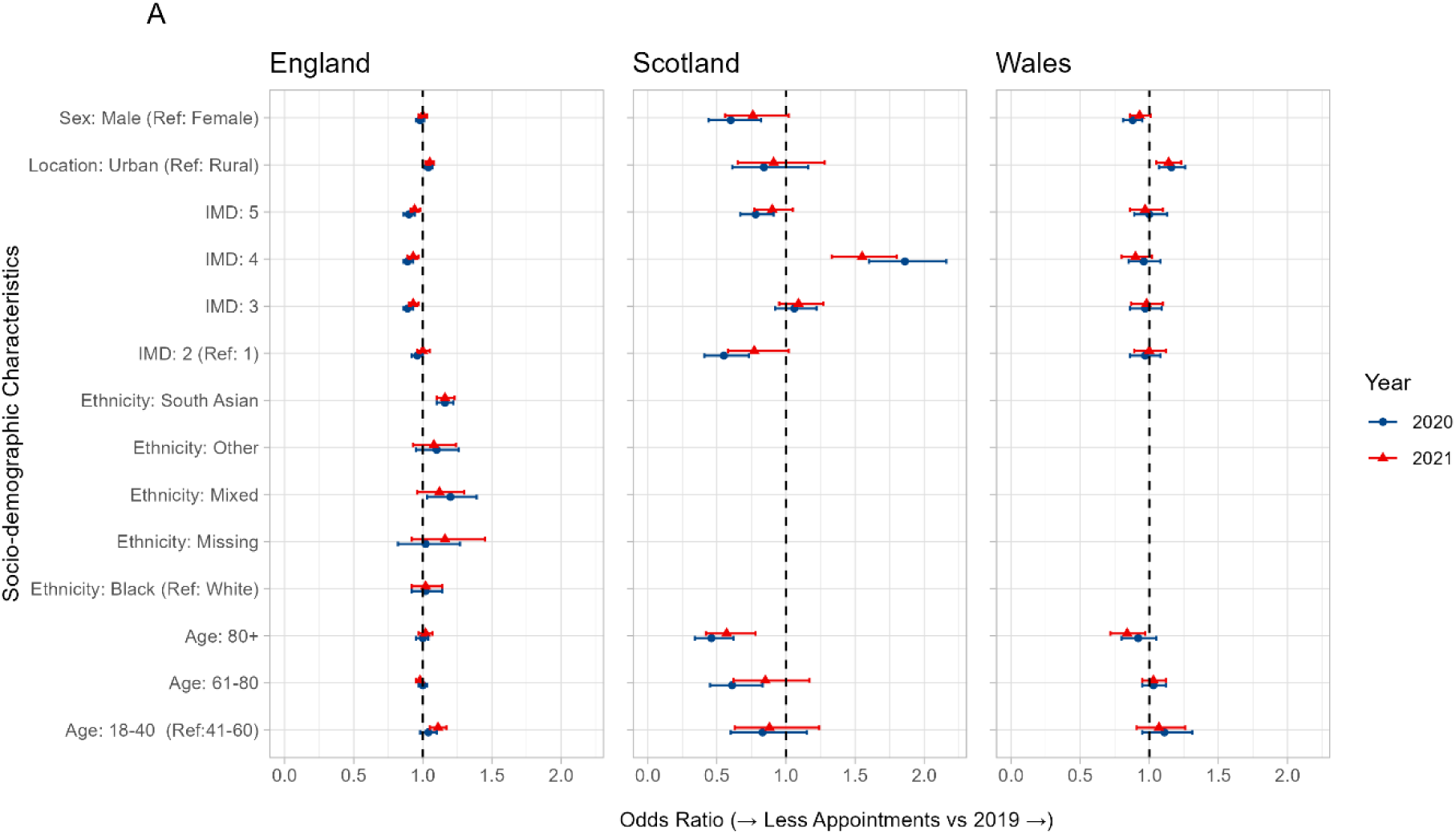
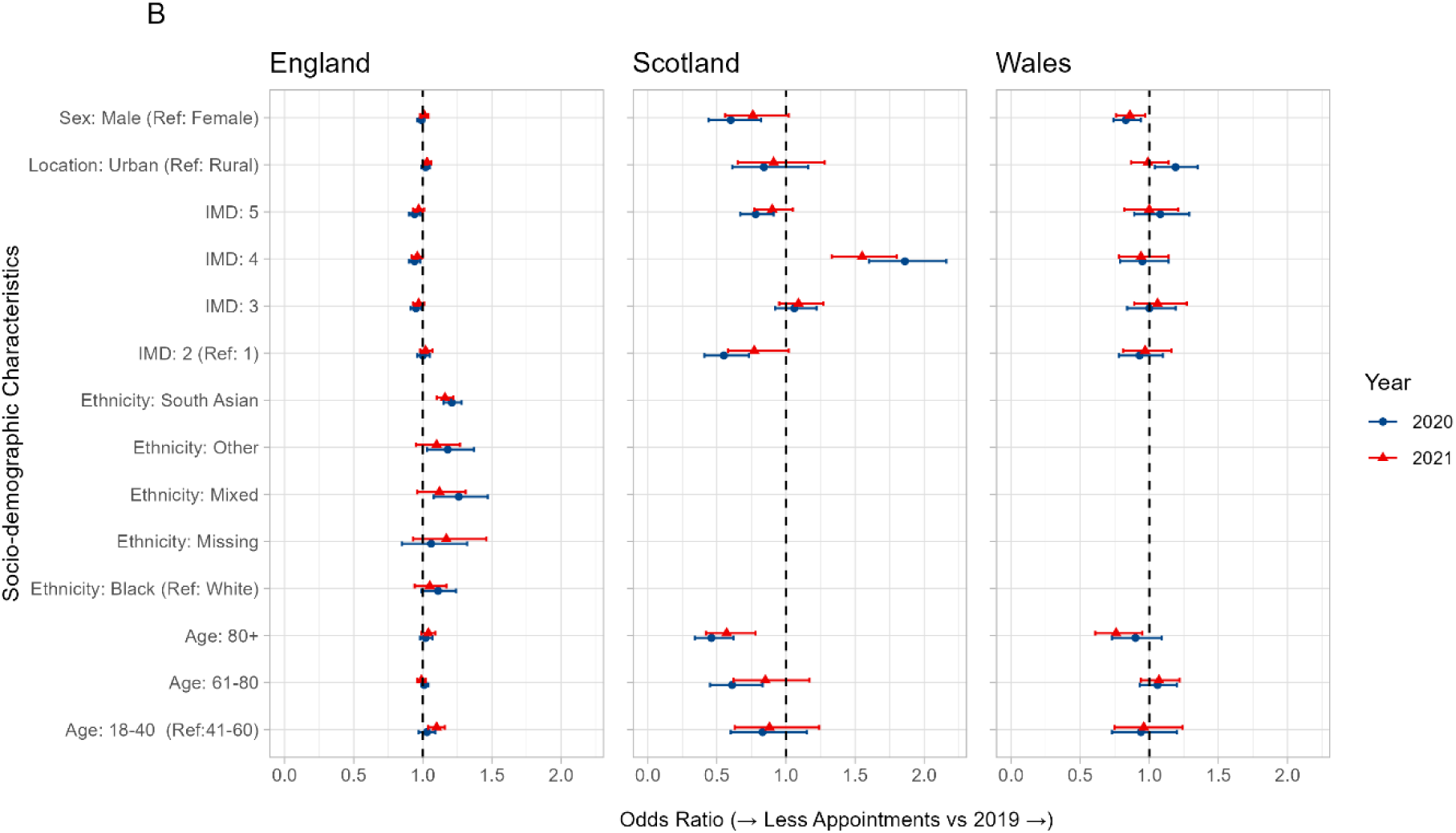
Forest plot of a) univariate and b) multivariate odds ratio and 95% confidence interval for having fewer rheumatology outpatient appointments in 2020 compared to 2019, by sociodemographic characteristic. An odds ratio above one indicates that people were more likely to have fewer appointments, and odds ratio below one indicates the people were less likely to have fewer appointments.

## Discussion

This is the first study to compare healthcare use and socio-demographic characteristics of people with RA in the three devolved nations of the UK during the COVID-19 pandemic and the recovery period. We found a reduction in the proportion of rheumatology outpatient appointments at the start of the pandemic in all three nations, however subsequent recovery varied. Since June 2020, in Wales, there has been a recovery in scheduled rheumatology appointments, albeit not to pre-pandemic levels. By contrast, in England and Scotland rheumatology outpatient appointments offered or attended had not recovered at the end of the study period. The proportion of patients with rheumatology appointments reduced over time into 2022 in Scotland and into 2023 in England. A high proportion of patients were not seen for several years, particularly in Scotland where 41% were not seen in 2019 and 2020. These people were frequently over 80 years old. In England, those of non-White ethnicity and those living in areas with higher deprivation had fewer appointments after the start of the pandemic.

This was a large study using electronic health record data from three nations, including for the first time primary care data from Scotland. This gives a unique insight into healthcare utilisation of an exemplar long-term condition requiring regular secondary care management across three different UK nations with devolved healthcare and different priorities and policies pre and post-COVID-19 pandemic. However, there are some limitations which need to be considered when interpreting findings; we did not have full population coverage in any data source which may affect representativeness, for example we only had data for two health boards in Scotland which are more rural and less deprived as measured by area-based measures of deprivation. However, we estimate that our primary care cohorts equate to 0.5%-0.8% prevalence of RA, which is in keeping with national estimates suggesting broad representativeness. The prevalence of rheumatoid arthritis in developed countries is estimated across studies to be approximately 0.5% to 1% of the adult population (22). In Wales and Scotland, analysis of hospital outpatient records described appointments scheduled, whereas in England the number of appointments attended were available. Whilst this needs to be taken into account when making comparisons, and may explain the higher number of appointments in Wales, overall, the non-attendance rates are low in rheumatology (23) and likely to be distributed evenly across the cohorts. There were no measures of disease severity in this data which meant we could not determine whether people with fewer appointments due to good disease control. Some information was unavailable, poorly documented or complex to include, such as smoking and ethnicity, which meant we could not fully investigate their impact across all countries.

Studies of the impact of the pandemic hospital service provision in people with RA have generally focused on 2020, and most have been survey based. Studies, in the UK, Europe, the US and Egypt have indicated that patients felt they had reduced access to care during the pandemic (11,24–28). In a survey of patients in the UK, only 4.5% had not seen a rheumatologist at all over 18 months, which is lower than our study, but may reflect selection bias in who completed the survey (25).

A Royal College of Physicians survey in July 2020, found that physicians across a range of specialities did not expect to be back at full operational capacity within 12 months, even without any further major pandemic events. Specifically, rheumatologists expected to be back to 75% capacity (29). Our results suggest that the recovery of rheumatology services has been slow. Potential reasons for the slow recovery of rheumatology services post COVID-19 pandemic are multi-factorial. Firstly, Rheumatology services were under considerable strain prior to the COVID-19 pandemic (4). Scotland has more rheumatology consultant and MDT vacancies, compared to Wales and England (4), which may explain why recovery is slower in Scotland. The COVID-19 pandemic put further strain on already stressed rheumatology services, where many rheumatologists were deployed to frontline COVID-19 care in the early months. As a result, waiting lists lengthened. Interestingly, this may be a specialty-specific finding, as our data showed that outpatient hospital appointments in other specialities have recovered to at least pre-pandemic levels, which is in line with findings from NHS England (30). However, including all specialties together may mean that specialties who have recovered well hide struggling specialties. For example, in contrast to rheumatology, cancer care was (understandably) prioritised during the pandemic and in the immediate post-pandemic time frame. As a result consultation rates for cancer increased during the initial lockdown, showing the impact of this policy (31). The observed changes may also partly reflect changes to the way services are delivered in Rheumatology, such as a shift towards patient initiated follow up whereby patients are not sent routine rheumatology clinic appointments.

A systematic review found that ethnic minorities, being of low social economic status and living a greater distance from the clinic was associated with not attending appointments (23).

Similarly a study of DMARD monitoring in primary care found the same groups missed monitoring, along with older patients (32). Although we did not measure non-attendance directly, the results are similar to our findings, where in England ethnic minorities and people living in more deprived areas had fewer outpatient hospital appointments. This is important as there is some evidence that ethnic minorities have worse outcomes in early RA (33).

Interestingly the systematic review found that rheumatology was one of the specialties with the lowest non-attendance rates.

### Implications

Rheumatic and musculoskeletal disease care has not been a priority to date, and we have seen a reduction in outpatient appointments within rheumatology, compared to a recovery of appointments in other specialties following the COVID-19 pandemic. People with RA would expect to see a member of the rheumatology multi-disciplinary team in clinic typically once a year as part of their overall disease management. The overall reduction in appointments we identified, and the large proportion of people who were not seen at all for a year or more may represent missed opportunities in RA management, potentially resulting in worse long-term outcomes (34).

Our study has also provided the opportunity for cross border learning in terms of data-driven approaches to service planning and access to primary and secondary care health data to support this, as well as evaluation of differing policy approaches across the devolved nations. We have identified groups of individuals (minority ethic groups and more deprived) that are more frequently not seen in rheumatology clinics. The wider RHEUMAPs study has used the data to create interactive geo-spatial maps in Wales to support local, regional and national service planning (35).

## Conclusions

Access to specialist rheumatology care for people with RA reduced at the start of the COVID-19 pandemic and had not recovered by the end of the study periods in England and Scotland; whilst Wales has shown some recovery. There are groups of patients who are missing out on specialist care: the elderly were frequently not seen at all over multiple years, and those of non-White ethnicity and those who were more deprived frequently had fewer outpatient hospital appointments. These may represent groups where there are missed opportunities for management of RA and potential for poorer long-term outcomes. This highlights the importance of tailored responses to different populations of the UK, particularly in times of, or response to significant healthcare stressors.

## Acknowledgements

We wish to thank Albasoft for access to primary care healthcare data in Scotland and the NHS Scotland electronic Data Research and Innovation Service Team (Public Health Scotland) for their involvement in obtaining approvals, provisioning, and linking data and the use of the secure analytical platform within the National Safe Haven. We are also very grateful for all the support received from the TPP Technical Operations team throughout this work, and for generous assistance from the information governance and database teams at NHS England and the NHS England Transformation Directorate. We would also like to thank Gary J Macfarlane and Ernest Choy (RHEUMAPs investigators) for their comments on the manuscript, Laura Moir (project administrator for RHEUMAPs) and Michelle Stevenson (patient partner on the RHEUMAPs study) for supporting recruitment of general practices in Scotland.

## Statements (funding, conflict of interest, ethics and data availability)

### Funding statement

In England, the OpenSAFELY platform is principally funded by grants from:

- NHS England [2023-2025];
- The Wellcome Trust (222097/Z/20/Z) [2020-2024];
- MRC (MR/V015737/1) [2020-2021].

Additional contributions to OpenSAFELY have been funded by grants from:

- MRC via the National Core Study programme, Longitudinal Health and Wellbeing strand (MC_PC_20030, MC_PC_20059) [2020-2022] and the Data and Connectivity strand (MC_PC_20058) [2021-2022];
- NIHR and MRC via the CONVALESCENCE programme (COV-LT-0009, MC_PC_20051) [2021-2024];
- NHS England via the Primary Care Medicines Analytics Unit [2021-2024].

The views expressed are those of the authors and not necessarily those of the NIHR, NHS England, UK Health Security Agency (UKHSA), the Department of Health and Social Care, or other funders. Funders had no role in the study design, collection, analysis, and interpretation of data; in the writing of the report; and in the decision to submit the article for publication.

Funding for the RHEUMAPS study and creation and analysis of the Scotland and Wales datasets used in this study was provided by the Nuffield Foundation Oliver Bird Fund (grant number OBF/44000.

LAT is funded by an NIHR Research Professorship NIHR302405.

### Ethics and information governance

In England, this study was approved by the Health Research Authority (REC reference 20/LO/0651) and by the LSHTM Ethics Board (reference 21863).

In Wales, the study was approved by the SAIL Information Governance Review Panel (approval number: 0419). All data used in this study can be accessed by request to SAIL.

Approvals for data linkage in Scotland were obtained from the Public Benefit and Privacy Panel for Health and Social Care, Scotland (reference number 1819-0286).

In England, NHS England is the data controller of the NHS England OpenSAFELY COVID-19 Service; TPP is the data processor; all study authors using OpenSAFELY have the approval of NHS England (36). This implementation of OpenSAFELY is hosted within the TPP environment which is accredited to the ISO 27001 information security standard and is NHS IG Toolkit compliant (37).

Patient data has been pseudonymised for analysis and linkage using industry standard cryptographic hashing techniques; all pseudonymised datasets transmitted for linkage onto OpenSAFELY are encrypted; access to the NHS England OpenSAFELY COVID-19 service is via a virtual private network (VPN) connection; the researchers hold contracts with NHS England and only access the platform to initiate database queries and statistical models; all database activity is logged; only aggregate statistical outputs leave the platform environment following best practice for anonymisation of results such as statistical disclosure control for low cell counts (1).

The service adheres to the obligations of the UK General Data Protection Regulation (UK GDPR) and the Data Protection Act 2018. The service previously operated under notices initially issued in February 2020 by the Secretary of State under Regulation 3(4) of the Health Service (Control of Patient Information) Regulations 2002 (COPI Regulations), which required organisations to process confidential patient information for COVID-19 purposes; this set aside the requirement for patient consent (2). As of 1 July 2023, the Secretary of State has requested that NHS England continue to operate the Service under the COVID-19 Directions 2020 (3). In some cases of data sharing, the common law duty of confidence is met using, for example, patient consent or support from the Health Research Authority Confidentiality Advisory Group (4).

Taken together, these provide the legal bases to link patient datasets using the service. GP practices, which provide access to the primary care data, are required to share relevant health information to support the public health response to the pandemic, and have been informed of how the service operates.

1. NHS Digital [Internet]. [cited 2023 Sep 20]. ISB1523: Anonymisation Standard for Publishing Health and Social Care Data. Available from: https://digital.nhs.uk/data-and-information/information-standards/information-standards-and-data-collections-including-extractions/publications-and-notifications/standards-and-collections/isb1523-anonymisation-standard-for-publishing-health-and-social-care-data
2. GOV.UK [Internet]. 2022 [cited 2023 Sep 20]. [Withdrawn] [withdrawn] Coronavirus (COVID-19): notice under regulation 3(4) of the Health Service (Control of Patient Information) Regulations 2002 – general. Available from: https://www.gov.uk/government/publications/coronavirus-covid-19-notification-of-data-controllers-to-share-information/coronavirus-covid-19-notice-under-regulation-34-of-the-health-service-control-of-patient-information-regulations-2002-general--2
3. NHS Digital [Internet]. [cited 2023 Sep 20]. COVID-19 Public Health Directions 2020. Available from: https://digital.nhs.uk/about-nhs-digital/corporate-information-and-documents/directions-and-data-provision-notices/secretary-of-state-directions/covid-19-public-health-directions-2020
4. Health Research Authority [Internet]. [cited 2023 Sep 20]. Confidentiality Advisory Group. Available from: https://www.hra.nhs.uk/about-us/committees-and-services/confidentiality-advisory-group/

### Data availability

In England, access to the underlying identifiable and potentially re-identifiable pseudonymised electronic health record data is tightly governed by various legislative and regulatory frameworks, and restricted by best practice. The data in the NHS England OpenSAFELY COVID-19 service is drawn from General Practice data across England where TPP is the data processor

TPP developers initiate an automated process to create pseudonymised records in the core OpenSAFELY database, which are copies of key structured data tables in the identifiable records. These pseudonymised records are linked onto key external data resources that have also been pseudonymised via SHA-512 one-way hashing of NHS numbers using a shared salt. University of Oxford, Bennett Institute for Applied Data Science developers and PIs, who hold contracts with NHS England, have access to the OpenSAFELY pseudonymised data tables to develop the OpenSAFELY tools.

These tools in turn enable researchers with OpenSAFELY data access agreements to write and execute code for data management and data analysis without direct access to the underlying raw pseudonymised patient data, and to review the outputs of this code. All code for the full data management pipeline — from raw data to completed results for this analysis — and for the OpenSAFELY platform as a whole is available for review at github.com/OpenSAFELY.

Access to NHS Scotland health data is governed by the NHS Scotland Public Benefit and Privacy Panel for Health and Social Care (HSC-PBPP). Access to the underlying pseudonymised health data used in this study is by application to the HSC-PBPP panel. All data used in this study can be accessed by request to SAIL.

The data management and analysis code for this paper was led by Ruth E Costello in England and Michael Parker in Wales and Scotland.

### Author contributions

SBacon: curated data (OpenSAFELY) and reviewed the manuscript. SBrophy conceptualised the study, acquired funding (RHEUMAPs study) , contributed to the methodology, reviewing and editing of the manuscript. REC conceptualised the study, contributed to the methodology, contributed to data visualisation and formal analysis, wrote the original draft, and reviewed and edited the manuscript. DE: curated data (OpenSAFELY) and reviewed the manuscript. BG: curated data (OpenSAFELY), acquired funding (CONVALESCENCE) and reviewed the manuscript. RJH conceptualised the study, acquired funding (RHEUMAPs study), contributed to the methodology, formal analysis, supervision, and writing, reviewing, and editing of the manuscript. JH conceptualised the study, contributed to the methodology, formal analysis, and writing, reviewing, and editing of the manuscript. JK contributed to the formal analysis, and reviewing and editing of the manuscript. BM: curated data (OpenSAFELY) and reviewed the manuscript. AM: curated data (OpenSAFELY) and reviewed the manuscript. MP contributed to the methodology, contributed to data visualisation and formal analysis, wrote the original draft, and reviewed and edited the manuscript. LT conceptualised the study, acquired funding (CONVALESCENCE), contributed to the methodology, supervision, reviewing and editing of the manuscript. All authors agreed to the final draft of the manuscript and publication.

