## Supplementary material for "Healthcare use in individuals with rheumatoid arthritis during the COVID-19 pandemic and beyond: A cohort study in three nations of the UK": Table S1, supplementary materials

### Contents

Supplementary Table S1 Number of other specialty outpatient appointments, and comparison to 2019, by nation and year. 2

Supplementary Table S2: Characteristics of patients by category of difference in the number of rheumatology outpatient appointments in 2020 compared with 2019 3

Supplementary Table S3: Characteristics of patients by category of difference in the number of rheumatology outpatient appointments in 2021 compared with 2019 5

Supplementary Table S4: Logistic regression results measuring characteristics of those who had fewer appointments in 2020 compared to 2019. 8

Supplementary Table S5: Logistic regression measuring characteristics of those who had fewer appointments in 2021 compared to 2019. 9

Supplementary Table S6: OpenSAFELY Collaborative 11

Supplementary Table S7: The LH&W NCS Collaborative 12

Supplementary text S8: RHEUMAPS study investigators 13

### Supplementary Table S1 Number of other specialty outpatient appointments, and comparison to 2019, by nation and year.

| | | April 2019-March 2020 | | | April 2020 – March 2021 | | | April 2020 – March 2021 | | | April 2022 - March 2023 | | |
|---|---|---|---|---|---|---|---|---|---|---|---|---|---|
| | | England | Wales | Scotland | England | Wales | Scotland | England | Wales | Scotland | England | Wales | Scotland |
| N* | | 138835 | 13021 | 3614 | 131195 | 12351 | 3412 | 124730 | 11791 | 3238 | 119295 | - | - |
| Number of All Appointments, n (column %) | Zero appointments | 20,570 (14.8) | 1,609 (12.36) | 933 (25.8) | 22,490 (17) | 2,278 (18.44) | 1384 (40.6) | 19,065 (15.2) | 2,042 (17.32) | 1256 (38.8) | 17,485 (14.7) | - | - |
| | 1-2 per year | 34,830 (25.1) | 3,584 (27.52) | 1401 (38.8) | 41,380 (31.3) | 4,144 (33.55) | 1166 (34.2) | 35,190 (28) | 3,156 (26.77) | 975 (30.1) | 32,645 (27.4) | - | - |
| | 3-5 per year | 35,655 (25.7) | 3,839 (29.48) | 844 (23.4) | 33,455 (25.3) | 3,452 (27.95) | 574 (16.8) | 31,410 (25) | 3,171 (26.89) | 615 (19.0) | 30,485 (25.6) | - | - |
| | 6 or more per year | 47,780 (34.4) | 3,989 (30.64) | 436 (12.1) | 34,750 (26.3) | 2,477 (20.06) | 288 (8.4) | 39,980 (31.8) | 3422 (29.02) | 392 (12.1) | 38,680 (32.4) | - | - |
| Difference in number of Appointments compared to 2019, n (column %) | Zero appointments both years | - | - | - | 10,415 (7.9) | 899 (7.3) | 641 (18.8) | 8,425 (617) | 725 (5.87) | 561 (17.3) | 7,280 (6.1) | - | - |
| | Fewer appointments | - | - | - | 65,930 (49.9) | 6,469 (54.8) | 1625 (47.6) | 56,480 (45.0) | 5,262 (42.6) | 1411 (43.6) | 52,895 (44.3) | - | - |
| | Same number or more appointments | - | - | - | 55,735 (42.2) | 4,983 (40.3) | 1146 (33.6) | 60,740 (48.3) | 5,804 (46.99) | 1266 (39.1) | 59,120 (49.6) | - | - |

* Counts for England rounded to the nearest 5.

### Supplementary Table S2: Characteristics of patients by category of difference in the number of rheumatology outpatient appointments in 2020 compared with 2019

| | | No appointments either year | | | Fewer appointments | | | Same number or more appointments | | |
|---|---|---|---|---|---|---|---|---|---|---|
| | | England | Wales | Scotland | England | Wales | Scotland | England | Wales | Scotland |
| | N | 28,895 | 2271 | 1403 | 45,530 | 4666 | 1139 | 57,580 | 5414 | 860 |
| Age category, n (row %) | 18 - 40 years | 1360 (18) | 116 (15.5) | 123 (48.4) | 2790 (36.9) | 298 (39.9) | 63 (24.8) | 3405 (45.1) | 332 (44.5) | 68 (26.8) |
| | 41 - 60 years | 6740 (17.7) | 662 (16.9) | 429 (38.8) | 13855 (36.3) | 1466 (37.4) | 392 (35.4) | 17570 (46) | 1793 (45.7) | 286 (25.8) |
| | 61 - 80 years | 14385 (20.8) | 1165 (17.7) | 663 (38.8) | 24145 (34.9) | 2514 (38.2) | 607 (35.5) | 30570 (44.2) | 2909 (44.2) | 439 (25.7) |
| | >80 years | 6410 (37.3) | 328 (29.9) | 188 (56.6) | 4740 (27.6) | 388 (35.4) | 77 (23.2) | 6035 (35.1) | 380 (34.7) | 67 (20.2) |
| Sex, n (row %) | Female | 19595 (20.9) | 1501 (17.3) | 873 (38.1) | 32790 (35) | 3360 (38.7) | 808 (35.3) | 41225 (44) | 3831 (44.1) | 610 (26.6) |
| | Male | 9300 (24.2) | 770 (21) | 530 (47.7) | 12740 (33.2) | 1306 (35.7) | 331 (29.8) | 16360 (42.6) | 1583 (43.3) | 250 (22.5) |
| Rural Urban classification, n (row %) | Rural | 7490 (22.9) | 990 (25.1) | 1008 (44.5) | 10960 (33.5) | 1395 (35.4) | 652 (28.8) | 14250 (43.6) | 1555 (39.5) | 607 (26.8) |
| | Urban | 21405 (21.6) | 1281 (15.2) | 395 (34.8) | 34565 (34.8) | 3271 (38.9) | 487 (42.9) | 43335 (43.6) | 3859 (45.9) | 253 (22.3) |
| IMD, n (row %) | 1 (Most deprived) | 5060 (21.6) | 378 (16) | 73 (34) | 8485 (36.2) | 907 (38.3) | 95 (44.2) | 9925 (42.3) | 1084 (45.8) | 47 (21.9) |

| | | | | | | | | | | |
|---|---|---|---|---|---|---|---|---|---|---|
| | 2 | 5785 (22.5) | 455 (16.7) | 187 (35.5) | 8975 (34.9) | 1025 (37.5) | 209 (39.7) | 10930 (42.5) | 1252 (45.8) | 131 (24.9) |
| | 3 | 6525 (21.9) | 555 (21.4) | 323 (34.1) | 10060 (33.8) | 973 (37.5) | 310 (32.7) | 13160 (44.2) | 1065 (41.1) | 315 (33.2) |
| | 4 | 5980 (21.6) | 521 (21.9) | 636 (52.7) | 9375 (33.9) | 885 (37.3) | 323 (26.8) | 12325 (44.5) | 968 (40.8) | 247 (20.5) |
| | 5 (Least deprived) | 5540 (21.8) | 362 (15.9) | 184 (36.4) | 8635 (34) | 876 (38.4) | 202 (39.9) | 11245 (44.2) | 1045 (45.8) | 120 (23.7) |
| smoking, n (row %) | Never smoked | 11055 (22.3) | 581 (18) | - | 17175 (34.7) | 1234 (38.3) | - | 21290 (43) | 1406 (43.7) | - |
| | Current smoker | 4120 (22) | 555 (19.6) | - | 6375 (34.1) | 1062 (37.5) | - | 8200 (43.9) | 1217 (42.9) | - |
| | Ex-smoker | 13635 (21.5) | 1135 (18) | - | 21855 (34.4) | 2370 (37.6) | - | 27970 (44.1) | 2791 (44.3) | - |
| | Unknown | 85 (25.8) | - | - | 120 (36.4) | - | - | 125 (37.9) | - | - |
| Time since first RA code, mean (standard deviation) | | 15 (42.9) | 10.52 (7.1) | 7.22 (4.63) | 10 (28.6) | 8.9 (5.34) | 7.74 (5.23) | 10 (28.6) | 9.29 (5.9) | 8 (5.18) |
| BMI, n (row %) | Missing | 4775 (26) | - | - | 5990 (32.6) | - | - | 7620 (41.4) | - | - |
| | Underweight | 645 (23.7) | - | - | 935 (34.4) | - | - | 1140 (41.9) | - | - |
| | Healthy range | 7175 (21) | - | - | 12025 (35.2) | - | - | 14995 (43.9) | - | - |
| | Overweight | 8270 (21.3) | - | - | 13480 (34.8) | - | - | 17035 (43.9) | - | - |
| | Obese | 6845 (21.3) | - | - | 11125 (34.7) | - | - | 14120 (44) | - | - |
| | Morbidly obese | 1180 (20.3) | - | - | 1975 (33.9) | - | - | 2670 (45.8) | - | - |

| | | | | | | | | | | |
|---|---|---|---|---|---|---|---|---|---|---|
| Ethnicity, n (row %) | White | 26125 (21.8) | - | - | 41135 (34.3) | - | - | 52610 (43.9) | - | - |
| | South Asian | 1505 (19.5) | - | - | 2960 (38.3) | - | - | 3270 (42.3) | - | - |
| | Black | 415 (23.6) | - | - | 595 (33.9) | - | - | 745 (42.5) | - | - |
| | Mixed | 155 (18.7) | - | - | 325 (39.2) | - | - | 350 (42.2) | - | - |
| | Other | 220 (22) | - | - | 360 (36) | - | - | 420 (42) | - | - |
| | Missing | 475 (59) | - | - | 145 (18) | - | - | 185 (23) | - | - |

### Supplementary Table S3: Characteristics of patients by category of difference in the number of rheumatology outpatient appointments in 2021 compared with 2019

| | | No appointments either year | | | Fewer appointments | | | Same number or more appointments | | |
|---|---|---|---|---|---|---|---|---|---|---|
| | | England | Wales | Scotland | England | Wales | Scotland | England | Wales | Scotland |
| | N | 26,480 | 2127 | 1348 | 44,650 | 2659 | 1006 | 54,445 | 5203 | 776 |
| Age category, n (row %) | 18 - 40 years | 1320 (17.8) | 110 (14.9) | 124 (50.6) | 2905 (39.2) | 291 (39.3) | 71 (29) | 3180 (39.2) | 339 (45.8) | 50 (20.4) |
| | 41 - 60 years | 6435 (17.2) | 609 (15.7) | 428 (39.7) | 14040 (37.4) | 1459 (37.7) | 378 (35.1) | 17045 (37.4) | 1805 (46.6) | 272 (25.2) |
| | 61 - 80 years | 13425 (20.3) | 1126 (17.9) | 632 (39) | 23445 (35.5) | 2408 (38.4) | 572 (35.3) | 29135 (35.5) | 2741 (43.7) | 417 (25.7) |
| | >80 years | 5300 (36.2) | 282 (31.2) | 164 (57.5) | 4260 (29.1) | 303 (33.6) | 84 (29.5) | 5085 (29.1) | 318 (35.2) | 37 (13) |
| Sex, n (row %) | Female | 17960 (20.1) | 1388 (21.3) | 852 (38.9) | 32205 (36) | 1388 (21.3) | 766 (35) | 39265 (36) | 3746 (57.4) | 573 (26.2) |
| | Male | 8520 (23.6) | 739 (21.3) | 496 (47.8) | 12440 (34.4) | 1271 (36.7) | 339 (32.7) | 15180 (34.4) | 1457 (42) | 203 (19.6) |

| | | | | | | | | | | |
|---|---|---|---|---|---|---|---|---|---|---|
| Rural Urban classification, n (row %) | Rural | 6940 (22.3) | 909 (24.1) | 983 (45.6) | 10680 (34.3) | 1348 (35.8) | 667 (30.9) | 13500 (34.3) | 1508 (40.1) | 508 (23.5) |
| | Urban | 19545 (20.7) | 1218 (15.2) | 365 (34.1) | 33965 (36) | 3113 (38.8) | 438 (40.9) | 40945 (36) | 3695 (46) | 268 (25) |
| IMD, n (row %) | 1 (Most deprived) | 4610 (20.7) | 366 (16.2) | 63 (31.5) | 8135 (36.6) | 870 (38.5) | 81 (40.5) | 9510 (36.6) | 1022 (45.3) | 56 (28) |
| | 2 | 5295 (21.7) | 421 (16.1) | 177 (35.8) | 8830 (36.2) | 1005 (38.5) | 186 (37.6) | 10285 (36.2) | 1182 (45.3) | 132 (26.7) |
| | 3 | 5940 (21) | 525 (21.1) | 303 (34) | 9920 (35) | 944 (38) | 327 (36.7) | 12450 (35) | 1014 (40.8) | 262 (29.4) |
| | 4 | 5525 (20.9) | 481 (21.1) | 624 (53.9) | 9225 (35) | 821 (36.1) | 325 (28.1) | 11630 (35) | 974 (42.8) | 208 (18) |
| | 5 (Least deprived) | 5110 (21.1) | 334 (15.4) | 181 (37.3) | 8540 (35.3) | 821 (37.9) | 186 (38.4) | 10570 (35.3) | 1011 (46.7) | 118 (24.3) |
| smoking, n (row %) | Never smoked | 10275 (21.6) | 566 (18.3) | - | 16835 (35.4) | 1197 (38.6) | - | 20475 (35.4) | 1336 (43.1) | - |
| | Current smoker | 3750 (21) | 494 (18.1) | - | 6290 (35.3) | 1034 (38) | - | 7775 (35.3) | 1194 (43.9) | - |
| | Ex-smoker | 12375 (20.7) | 1067 (17.9) | - | 21395 (35.7) | 2230 (37.4) | - | 26085 (35.7) | 2673 (44.8) | - |
| | Unknown | 85 (26.6) | - | - | 125 (39.1) | - | - | 110 (39.1) | - | - |
| Time since first RA code, mean (standard deviation) | | 15 (42.9) | 10.55 (7.01) | 7.25 (4.62) | 10 (28.6) | 8.91 (10.40) | 7.84 (5.4) | 10 (28.6) | 9.13 (5.83) | 7.79 (4.93) |

| | | | | | | | | | | |
|---|---|---|---|---|---|---|---|---|---|---|
| BMI, n (row %) | Missing | 4320 (24.7) | - | - | 6045 (34.6) | - | - | 7125 (34.6) | - | - |
| | Underweight | 505 (21.1) | - | - | 880 (36.8) | - | - | 1005 (36.8) | - | - |
| | Healthy range | 6530 (20.2) | - | - | 11550 (35.7) | - | - | 14230 (35.7) | - | - |
| | Overweight | 7680 (20.7) | - | - | 13210 (35.6) | - | - | 16180 (35.6) | - | - |
| | Obese | 6350 (20.7) | - | - | 10970 (35.7) | - | - | 13410 (35.7) | - | - |
| | Morbidly obese | 1100 (19.7) | - | - | 1990 (35.6) | - | - | 2495 (35.6) | - | - |
| Ethnicity, n (row %) | White | 23920 (21) | - | - | 40270 (35.4) | - | - | 49670 (35.4) | - | - |
| | South Asian | 1430 (19) | - | - | 2955 (39.3) | - | - | 3135 (39.3) | - | - |
| | Black | 370 (21.8) | - | - | 600 (35.4) | - | - | 725 (35.4) | - | - |
| | Mixed | 140 (17.4) | - | - | 315 (39.1) | - | - | 350 (39.1) | - | - |
| | Other | 195 (20.4) | - | - | 355 (37.2) | - | - | 405 (37.2) | - | - |
| | Missing | 425 (57.8) | - | - | 150 (20.4) | - | - | 160 (20.4) | - | - |

### Supplementary Table S4: Logistic regression results measuring characteristics of those who had fewer appointments in 2020 compared to 2019.

| | | Univariable, odds ratio (95% confidence intervals) | | | Multivariable, odds ratio (95% confidence intervals) | | |
|---|---|---|---|---|---|---|---|
| **Variable** | **Group** | **England** | **Scotland** | **Wales** | **England** | **Scotland** | **Wales** |
| Age | 18-40 | 1.04 (0.98, 1.1) | 0.83(0.6, 1.15) | 1.11(0.95, 1.31) | 1.03 (0.97, 1.09) | 0.62(0.45, 0.85) | 0.94(0.73, 1.2) |
| | 41-60 | Reference | Reference | Reference | Reference | Reference | Reference |
| | 61-80 | 1 (0.97, 1.03) | 0.61(0.45, 0.83) | 1.03(0.95, 1.12) | 1.01 (0.99, 1.04) | 0.99(0.84, 1.16) | 1.06(0.93, 1.2) |
| | >80 | 1 (0.95, 1.04) | 0.46(0.34, 0.62) | 0.92(0.8, 1.05) | 1.02 (0.98, 1.07) | 0.52(0.39, 0.7) | 0.9(0.73, 1.09) |
| Sex | Male | 0.98 (0.95, 1.01) | 0.6(0.44, 0.82) | 0.88(0.81, 0.95) | 0.99 (0.96, 1.01) | 0.81(0.69, 0.95) | 0.83(0.74, 0.94) |
| Rural-urban classification | Urban | 1.04 (1.01, 1.07) | 0.84(0.61, 1.16) | 1.16(1.07, 1.26) | 1.02 (0.99, 1.05) | 1.7(1.44, 2) | 1.19(1.04, 1.35) |
| IMD | 1 (Most deprived) | Reference | Reference | Reference | Reference | Reference | Reference |
| | 2 | 0.96 (0.92, 1) | 0.55(0.41, 0.73) | 0.97(0.86, 1.08) | 1 (0.96, 1.05) | 0.91(0.66, 1.26) | 0.93(0.78, 1.1) |
| | 3 | 0.89 (0.86, 0.93) | 1.06(0.92, 1.22) | 0.97(0.86, 1.09) | 0.95 (0.91, 0.99) | 0.77(0.56, 1.06) | 1(0.84, 1.19) |
| | 4 | 0.89 (0.86, 0.93) | 1.86(1.6, 2.16) | 0.96(0.85, 1.08) | 0.94 (0.9, 0.98) | 0.61(0.44, 0.83) | 0.95(0.79, 1.14) |
| | 5 (Least deprived) | 0.9 (0.86, 0.94) | 0.78(0.67, 0.91) | 1(0.89, 1.13) | 0.94 (0.9, 0.98) | 0.88(0.63, 1.22) | 1.08(0.89, 1.29) |
| Ethnicity | White | Reference | Reference | Reference | Reference | Reference | Reference |
| | South Asian | 1.16 (1.1, 1.22) | - | - | 1.21 (1.15, 1.28) | - | - |
| | Black | 1.02 (0.92, 1.14) | - | - | 1.11 (0.99, 1.24) | - | - |

| | | | | | | | |
|---|---|---|---|---|---|---|---|
| | Mixed | 1.2 (1.03, 1.39) | - | - | 1.26 (1.08, 1.47) | - | - |
| | Other | 1.1 (0.95, 1.26) | - | - | 1.18 (1.03, 1.37) | - | - |
| | Missing | 1.02 (0.82, 1.27) | - | - | 1.06 (0.85, 1.32) | - | - |

### Supplementary Table S5: Logistic regression measuring characteristics of those who had fewer appointments in 2021 compared to 2019.

| | | **Univariable, odds ratio (95% confidence intervals)** | | | **Multivariable, odds ratio (95% confidence intervals)** | | |
|---|---|---|---|---|---|---|---|
| **Variable** | **Group** | **England** | **Scotland** | **Wales** | **England** | **Scotland** | **Wales** |
| Age | 18-40 | 1.11 (1.05, 1.17) | 0.88(0.63, 1.24) | 1.07(0.91, 1.26) | 1.1 (1.04, 1.16) | 0.79(0.58, 1.06) | 0.96(0.75, 1.24) |
| | 41-60 | Reference | Reference | Reference | Reference | Reference | Reference |
| | 61-80 | 0.98 (0.95, 1) | 0.85(0.62, 1.17) | 1.03(0.95, 1.12) | 0.99 (0.96, 1.02) | 0.99(0.84, 1.17) | 1.07(0.94, 1.22) |
| | >80 | 1.02 (0.97, 1.07) | 0.57(0.42, 0.78) | 0.84(0.72, 0.97) | 1.04 (0.99, 1.09) | 0.75(0.56, 1) | 0.76(0.61, 0.95) |
| Sex | Male | 1 (0.97, 1.03) | 0.76(0.56, 1.02) | 0.93(0.86, 1.01) | 1.01 (0.98, 1.04) | 0.93(0.8, 1.1) | 0.86(0.76, 0.97) |
| Rural-urban classification | Urban | 1.05 (1.02, 1.08) | 0.91(0.65, 1.28) | 1.14(1.05, 1.23) | 1.03 (1, 1.06) | 1.49(1.26, 1.76) | 0.99(0.87, 1.14) |
| IMD | 1 (Most deprived) | Reference | Reference | Reference | Reference | Reference | Reference |
| | 2 | 1 (0.96, 1.05) | 0.77(0.58, 1.02) | 1(0.89, 1.12) | 1.02 (0.98, 1.07) | 0.95(0.68, 1.34) | 0.97(0.81, 1.16) |
| | 3 | 0.93 (0.9, 0.97) | 1.09(0.95, 1.27) | 0.98(0.87, 1.1) | 0.97 (0.93, 1.01) | 1.01(0.73, 1.41) | 1.06(0.89, 1.27) |

| | | | | | | | |
|---|---|---|---|---|---|---|---|
| | 4 | 0.93 (0.89, 0.97) | 1.55(1.33, 1.8) | 0.9(0.8, 1.02) | 0.96 (0.92, 1) | 0.71(0.51, 0.98) | 0.94(0.78, 1.14) |
| | 5 (Least deprived) | 0.94 (0.91, 0.98) | 0.9(0.77, 1.05) | 0.97(0.86, 1.1) | 0.97 (0.93, 1.01) | 0.95(0.68, 1.35) | 1(0.82, 1.21) |
| Ethnicity | White | Reference | Reference | Reference | Reference | Reference | Reference |
| | South Asian | 1.16 (1.1, 1.23) | - | - | 1.16 (1.1, 1.22) | - | - |
| | Black | 1.02 (0.92, 1.14) | - | - | 1.05 (0.94, 1.17) | - | - |
| | Mixed | 1.12 (0.96, 1.3) | - | - | 1.12 (0.96, 1.31) | - | - |
| | Other | 1.08 (0.93, 1.24) | - | - | 1.1 (0.95, 1.27) | - | - |
| | Missing | 1.11 (1.05, 1.17) | 0.88(0.63, 1.24) | 1.07(0.91, 1.26) | 1.17 (0.93, 1.46) | - | - |

### Supplementary Table S6: OpenSAFELY Collaborative

| First name and middle initial | Surname |
|---|---|
| Alex J | Walker |
| Brian | MacKenna |
| Peter | Inglesby |
| Ben | Goldacre |
| Helen J | Curtis |
| Caroline E | Morton |
| Jessica | Morley |
| Amir | Mehrkar |
| Sebastian CJ | Bacon |
| George | Hickman |
| Richard | Croker |
| David | Evans |
| Tom | Ward |
| Nicholas J | DeVito |
| Louis | Fisher |
| Amelia CA | Green |
| Jon | Massey |
| Rebecca M | Smith |
| William J | Hulme |
| Simon | Davy |
| Colm D | Andrews |
| Lisa EM | Hopcroft |
| Henry | Drysdale |
| Iain | Dillingham |
| Robin Y | Park |
| Rose | Higgins |
| Christine | Cunningham |
| Milan | Wiedemann |
| Linda | Nab |
| Steven | Maude |
| Orla | Macdonald |
| Ben FC | Butler-Cole |
| Thomas | O'Dwyer |
| Catherine L | Stables |
| Christopher | Wood |
| Andrew D | Brown |
| Victoria | Speed |
| Lucy | Bridges |
| Andrea L | Schaffer |
| Caroline E | Walters |
| Christopher T | Rentsch |

| | |
|---|---|
| Krishnan | Bhaskaran |
| Anna | Schultze |
| Elizabeth J | Williamson |
| Helen I | McDonald |
| Laurie A | Tomlinson |
| Rohini | Mathur |
| Rosalind M | Eggo |
| Kevin | Wing |
| Angel YS | Wong |
| John | Tazare |
| Richard | Grieve |
| Daniel J | Grint |
| Sinead | Langan |
| Kathryn E | Mansfield |
| Ian J | Douglas |
| Stephen JW | Evans |
| Liam | Smeeth |
| Jemma L | Walker |
| Viyaasan | Mahalingasivam |
| Harriet | Forbes |
| Thomas E | Cowling |
| Emily L | Herrett |
| Ruth E | Costello |
| Bang | Zheng |
| Edward P K | Parker |
| Christopher | Bates |
| Jonathan | Cockburn |
| John | Parry |
| Frank | Hester |
| Sam | Harper |
| Shaun | O'Hanlon |
| Alex | Eavis |
| Richard | Jarvis |
| Dima | Avramov |
| Paul | Griffiths |
| Aaron | Fowles |
| Nasreen | Parkes |
| Brian | Nicholson |
| Rafael | Perera |
| David | Harrison |
| Kamlesh | Khunti |
| Jonathan AC | Sterne |
| Jennifer | Quint |

### Supplementary Table S7: The LH&W NCS Collaborative

| First name | Surname |
|---|---|
| Nishi | Chaturvedi |
| Chloe | Park |
| Alisia | Carnemolla |
| Dylan | Williams |
| Anika | Knueppel |
| Andy | Boyd |
| Emma L | Turner |
| Katharine M | Evans |
| Richard | Thomas |
| Samantha | Berman |
| Stela | McLachlan |
| Matthew | Crane |
| Rebecca | Whitehorn |
| Jacqui | Oakley |
| Diane | Foster |
| Hannah | Woodward |
| Kirsteen C | Campbell |
| Nicholas | Timpson |
| Alex | Kwong |
| Ana Goncalves | Soares |
| Gareth | Griffith |
| Renin | Toms |
| Louise | Jones |
| Herbert, | Annie |
| Ruth | Mitchell |
| Tom | Palmer |
| Jonathan | Sterne |
| Venexia | Walker |
| Lizzie | Huntley |
| Laura | Fox |
| Rachel | Denholm |
| Rochelle | Knight |
| Kate | Northstone |
| Arun | Kanagaratnam |
| Elsie | Horne |
| Harriet | forbes |
| Teri | North |
| Kurt | Taylor |
| Marwa AL | Arab |
| Scott | Walker |
| Jose IC | Coronado |
| Arun S | Karthikeyan |
| George | Ploubidis |
| Bettina | Moltrecht |
| Charlotte | Booth |
| Sam | Parsons |
| Bozena | Wielgoszewska |
| Charis | Bridger-Staatz |
| Claire | Steves |
| Ellen | Thompson |
| Paz | Garcia |
| Nathan | Cheetham |
| Ruth | Bowyer |
| Maxim | Freydin |
| Amy | Roberts |
| Ben | Goldacre |
| Alex | Walker |
| Jess | Morley |
| William | Hulme |
| Linda | Nab |
| Louis | Fisher |
| Brian | MacKenna |
| Colm | Andrews |
| Helen | Curtis |
| Lisa | Hopcroft |
| Amelia | Green |
| Praveetha | Patalay |
| Jane | Maddock |
| Kishan | Patel |
| Jean | Stafford |
| Wels | Jacques |
| Kate | Tilling |
| John | Macleod |
| Eoin | McElroy |
| Anoop | Shah |
| Richard | Silverwood |
| Spiros | Denaxas |
| Robin | Flaig |
| Daniel | McCartney |
| Archie | Campbell |
| Laurie | Tomlinson |
| John | Tazare |
| Bang | Zheng |
| Liam | Smeeth |
| Emily | Herrett |
| Thomas | Cowling |
| Kate | Mansfield |
| Ruth E | Costello |
| Kevin | Wang |
| Kathryn | Mansfield |
| Viyaasan | Mahalingasivam |
| Ian | Douglas |
| Sinead | Langan |
| Sinead | Brophy |
| Michael | Parker |
| Jonathan | Kennedy |
| Rosie | McEachan |
| John | Wright |
| Kathryn | Willan |
| Ellena | Badrick |
| Gillian | Santorelli |
| Tiffany | Yang |
| Bo | Hou |
| Andrew | Steptoe |
| Di Gessa, | Giorgio |
| Jingmin | Zhu |
| Paola | Zaninotto |
| Angela | Wood |
| Genevieve | Cezard |
| Samantha | Ip |
| Tom | Bolton |
| Alexia | Sampri |
| Elena | Rafeti |
| Fatima | Almaghrabi |
| Aziz | Sheikh |
| Syed A | Shah |
| Vittal | Katikireddi |
| Richard | Shaw |
| Olivia | Hamilton |
| Michael | Green |
| Theocharis | Kromydas |
| Daniel | Kopasker |
| Felix | Greaves |
| Robert | Willans |
| Fiona | Glen |
| Steve | Sharp |
| Alun | Hughes |
| Andrew | Wong |
| Lee Hamill | Howes |
| Alicja | Rapala |
| Lidia | Nigrelli |
| Fintan | McArdle |
| Chelsea | Beckford |
| Betty | Raman |
| Richard | Dobson |
| Amos | Folarin |
| Callum | Stewart |
| Yatharth | Ranjan |
| Jd | Carpentieri |
| Laura | Sheard |
| Chao | Fang |
| Sarah | Baz |
| Andy | Gibson |
| John | Kellas |
| Stefan | Neubauer |
| Stefan | Piechnik |
| Elena | Lukaschuk |
| Laura C | Saunders |
| James M | Wild |
| Stephen | Smith |
| Peter | Jezzard |
| Elizabeth | Tunnicliffe |
| Zeena-Britt | Sanders |
| Lucy | Finnigan |
| Vanessa | Ferreira |
| Mark | Green |
| Rebecca | Rhead |
| Milla | Kibble |
| Yinghui | Wei |
| Agnieszka | Lemanska |
| Francisco | Perez-Reche |
| Dominik | Piehlmaier |
| Lucy | Teece |
| Edward | Parker |

### Supplementary text S8: RHEUMAPS study investigators

The RHEUMAPS study investigators includes: Rosemary J Hollick (Chief Investigator) (Senior Clinical Lecturer, University of Aberdeen, Honorary Consultant Rheumatologist, NHS Grampian), Corri Black (Emeritus Professor, University of Aberdeen), Sinead Brophy (Professor of Health Data Science, Swansea University), Ernest Choy (Head of Rheumatology and Translational Research, Cardiff University), Gary Macfarlane (Clinical Chair in Epidemiology, University of Aberdeen), Louise Bennett (University of Glasgow), Lorna Philip (Professor of Geography and Environment, University of Aberdeen), Michelle Stevenson (Patient Partner), Denise McFarlane (GP, NHS Grampian and Chair, External Advisory Group), Laura Moir (Study Coordinator, University of Aberdeen) and Public Contributors Ian Allotay, Philip Bell, Amanda Cheesley, Charlotte Marlow, Farzana Kausir, Emily Lam and Inga Wood.
